# Predictor of neoplasms and body composition with machine learning models

**DOI:** 10.1101/2023.05.07.23289621

**Authors:** Mª Jesús Fuentes Sebio

## Abstract

**Background:** The tissue microenvironment of neoplastic diseases differs from that of normal cells. Their extracellular matrix, innervation, metabolism, as well as interstitial fluid and intercellular interconnections mark clear physical differences between normal and cancerous cellular ecosystems. Detecting these physical changes early without using diagnostic methods that are harmful and uncomfortable for the patient is a major challenge for the medical-scientific community. Electrical bioimpedance supported by machine learning techniques can provide clues to incipient preneoplastic tissue changes.

**Methods:** In this study, 7 predictive machine learning models were developed using a database with bioimpedanciometric data from a group of healthy individuals and another group of patients who had or were suffering from cancer at the time of measurement.

**Results:** The *Random Forest* was the model that reported the best Accuracy, reaching over 90% of hits.

**Conclusions:** These results open the door to future research linking changes in body composition and pretumoral tissue environments using machine learning tools.

## 1 Introduction

Cancer encompasses a group of diseases characterized by the abnormal and uncontrolled division of anarchic cells that infiltrate and destroy normal tissues [56]. Its development requires a three-dimensional microenvironment that favors it, in which mechanical forces as well as biomolecular gradients and non-organic components come into play [52]. Neoplastic cells have an impressive capacity to evade therapy directed against them, and it is necessary to find medical clues indicating the development of favorable conditions for their development.

The study of the electrical properties of the organism to estimate body composition in both healthy and diseased individuals has been used for years with **bioimpedancemetry** (BIA) [53, 54], one of the most widely used methods to analyze body composition due to its ease of use, safety, reproducibility, low cost and good accuracy [29], although without reaching the precision of other more sophisticated techniques such as Computerized Axial Tomography (CT), Magnetic Resonance Imaging, Dual X Absorptiometry [9], Hydrometry, Isotope Dilution, etc., whose use is limited by their cost and side effects [4][18]. Bioimpedance can be used at any age and, except in some situations that contraindicate it, such as the use of pacemakers, it is a safe technique.

The physical basis of BIA is based on the resistance of the different body structures to the passage of a low-voltage electric current [11, 10]. This means that if we modify the electrical frequency or the physical-chemical conditions of the organism, the resistance to the passage of the current also changes [8]. Based on these properties, we define the **conductance** as the facility of a material to allow the flow of electric current and, on the contrary, the **resistance** is the difficulty to the passage of the same one.

There is no doubt that our organism is not a simple [48] circuit and as such, other physical components come into play, as is the case of the **capacitors**, whose function is to store energy to release it later. Our body capacitors are the cell membranes and interfaces, thus, we define the **electrical capacitance** as their capacity to store energy and the **reactance**, the resistance offered by these cell capacitors to the passage of electric current.

The **resistance** and the **reactance**, that together we call **Impedance (Z)**, are the factors that oppose resistance to the passage of electric current (Figure 1). The values of the Impedance depend on the voltage and the intensity of the electric current, as we can see in Figure 2.

**Figure 1:**
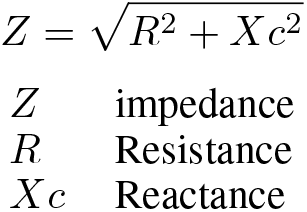
Corporal impedance.

**Figure 2:**
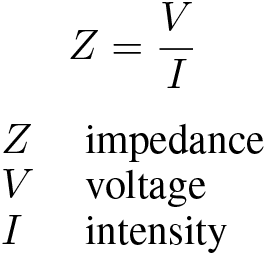
Impedance formula.

Bioimpedance is a simple and reliable method to identify groups of individuals with alterations in the quantity and distribution of body fluids as well as to assess the state of cell membranes [13]. The most commonly used equipment today is in the frequency range of 1 kHz - 1 MHz. The electric current flows through the extracellular water at low frequencies (1 - 5 kHz), which determines that the reactance is minimal [70]. If we increase the frequency (50 - 100 kHz), electricity penetrates into the intercellular space, but the membranes hinder it by acting as capacitors. If the frequency continues to rise, electricity flows through the extra and intracellular water as well as through the intervening compartments [39].

The angle formed by the above three vectors is called the “phase angle” (Figure 3) [12, 22, 21] and is one of the most widely used parameters in research. The **phase angle** shows a positive relationship with the integrity and optimal functionality of the body cells (reflected in the reactance) and is negatively associated with the degree of hydration of the various tissues (resistance) [47, 48].

**Figure 3:**
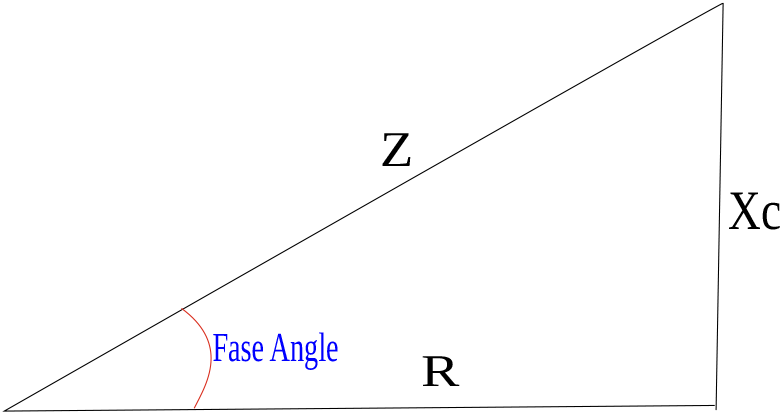
Relations between the components of bioimpedance.

To obtain the value of the phase angle we have to apply the Formula 1 [26]:

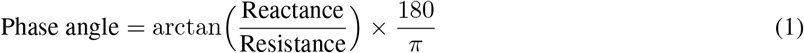

It is usually calculated with frequencies of 50 kHz on the right side of the body, however multi-frequency equipment calculates it on both sides [60].

The phase angle value reflects the integrity of cell membranes as well as intracellular and extracellular water concentrations. Low values indicate damage, destructuring of the cell wall and even cell death [14, 15, 20], being considered very useful to evaluate inflammatory abnormalities [69, 27]. Low phase angle values are a good prognostic marker in cardiac, renal, hepatic, infectious pathologies and cancer [51, 55], among others [47]. They give us clues to poor clinical evolution in individuals with cancer [59, 33], speculating that the alteration of the corporal homeostasis as well as an increase of inflammatory cytokines constitute the underlying basis [36, 37, 38]. High values, on the contrary, reflect optimal cellular integrity.

In the scientific literature, several research papers used impedance-based techniques to distinguish tumors from healthy tissues [30, 31, 32]. Anarchic and uncontrolled cell proliferation as well as a misconfiguration of the cell stroma at the tumor level results in alterations of the electrical charges; in fact, the most aggressive tumors entail cellular alterations at the systemic level that reduce survival [35].

Detecting minimal precancerous tissue changes is a major challenge for the scientific community, and new artificial intelligence tools are a breakthrough in discovering patterns and correlations in a data set in order to make predictive analyses of the data, much in the same way as human reasoning. **Machine learning** learns from the data, from a training dataset and generates a predictive model [23, 58], thus improving statistical analysis [57].

## 2 Objectives

The aim of this research work is to predict with machine learning algorithms whether a person, object of the study, is in a clinical situation with a high probability of suffering a neoplastic degeneration based on bioimpedanciometric studies.

## 3 Methodology

Observational, retrospective, cross-sectional study including bioimpedanciometric records of 912 healthy individuals and 1139 records of 88 individuals who suffers or suffered some type of neoplasms at the time of analysis, attended at Dra. Fuentes Medical Nutrition Center’s office in the period between 2015 and 2022. Pregnant women and individuals under 18 years of age were excluded. The multi-frequency BIA equipment TANITA MC-980MA with 8 electrodes [28] was the bioimpedance meter at 1, 5, 50, 250, 500 and 1000 kHz frequency.

The design and training of the neural network was carried out using the open source programming language **R** using the package **caret** (**c**lassification **a**nd **r**egression **t**raining), developed by *Max Kuhn* [46]. This interface has tools for the development of predictive models based on classification and regression processes and it is available at http://CRAN.R-project.org/package=caret.

The evaluation of the quality of the algorithm when making predictions is carried out with two metrics: *Accuracy* and *Kappa*. The former reports the percentage of correctly predicted observations and the *Kappa*, or *Cohen’s Kappa metric*, is *Accuracy’s* standardized result for accurate random results.

The machine learning models used were:

- **k-Nearest Neighbors (knn)**: Supervised learning method that classifies new data based on the distances of nearest neighbor values [42].
- **Logistic regression**: Used to model data that does not fit the linear regression model. Calculates the probability of a binary qualitative variable as a function of different quantitative variables [72].
- **Linear Discriminant Analysis**: Supervised classification method in which groups are organized according to their characteristics [72].
- **Single C5.0 Tree**: Model made up of simple classification trees [74].
- **Random Forest**: This algorithm created in 2001 by Leo Breiman uses multiple decision trees, training different parts of them, thus reducing the variance [45]. It has great flexibility and optimal predictive results, which is why it is widely used [50].
- **Stochastic Gradient Boosting**: It is based on the construction of several decision trees with random data from subsets of the data [74].
- **Neural Networks**: Inspired by the structure and functioning of our brain neural networks [45].

## 4 Exploratory data analysis

The participants were divided into healthy and pathological: 912 records from healthy individuals and 1139 bioimpedance measurements from 89 pathological individuals. The smaller number of pathological individuals and the need for machine learning to be able to recognize as many “unhealthy” individuals as possible for learning, motivated the selection of the records in this way.

The types of tumors are specified in the Table 1, with breast, uterus and thyroid neoplasms accounting for the largest number of individuals. Exploratory analysis of the data reflects a lower phase angle value in individuals in the pathologic group compared to individuals in the normal group 10. A reduced phase angle value reflects an alteration in tissue electrical properties [59], which suggests an unfavorable evolution of their health status

**Table 1.**

| *Neoplasm* | *No. Cases* |
| --- | --- |
| Breast | 33 |
| Thyroid | 12 |
| Uterus | 11 |
| Melanoma | 8 |
| Colon | 5 |
| Lymphoma | 5 |
| Lung | 3 |
| Pancreas | 3 |
| Ovary | 2 |
| Prostate | 2 |
| Bladder | 1 |
| Bone | 1 |
| Kidney | 1 |
| Testicle | 1 |

Individuals in the pathological group have a higher Body Mass Index (BMI), as can be seen in Figures 9, 10 and 11. This confirms the relationship between excess body mass and the development of different types of cancer reported in analyses conducted by the World Cancer Research Fund (WCRF) [77, 78]. What is most striking from the exploratory analysis of the data is that normally phase angle declines with age in healthy individuals. However, this does not occur in individuals in the pathological group in which the relationship between phase angle and age is not linear (Figures 12 and 13.

## 5 Training of machine learning models

The various machine learning algorithms were applied to the training data in order to achieve the best result.

### 5.1 k-Nearest Neighbors (kNN)

This is a very simple algorithm that attempts to identify similar observations. It uses k as the only parameter that tells us the number of neighboring observations used. In this case the values of k that were included were: 1, 2, 5, 10, 15, 20, 30 and 50. The highest Accuracy was achieved with k = 1, as we can see in the result:

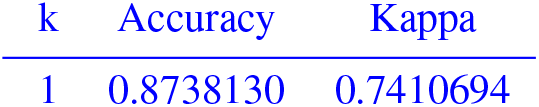

### 5.2 Logistic Regression

Logistic regression is a type of regression in which the dependent variable is categorical. It does not use hyperparameters and is a good technique for estimating probabilities[75].

The best training result was:

| Accuracy | Kappa |
| --- | --- |
| 0.8221619 | 0.6388024 |

### 5.3 Linear Discriminant Analysis

Machine learning classification method based on Bayes’ theorem. It calculates the probability that an observation belongs to one of the groups to be studied. It does not use any hyperparameter.

The result obtained was:

| Accuracy | Kappa |
| --- | --- |
| 0.8186268 | 0.6307069 |

### 5.4 Single C5.0 Tree

Developed by J.Ross Quinlan [75], this algorithm is simple but very efficient with the ability to exclude unhelpful features [75] and is very productive even on large data sets. It does not need hyperparameters.

Final result taking into account the best Accuracy:

| Accuracy | Kappa |
| --- | --- |
| 0.8774582 | 0.7499857 |

### 5.5 Random Forest

This method, although also simple, is capable of processing tables with unbalanced data [23]. Its algorithm uses the parameters: **mtry** and **min.node.size**, the former representing the number of randomly selected predictor variables in each tree and the parameter “min.node.size” setting the minimum size of the node to be split.

The hyperparameters used were mtry = c(3, 4, 5, 7) and min.node.size = c(2, 3, 4, 5, 10, 15, 20, 30). Final result taking into account the best Accuracy:

| mtry | min.node.size | Accuracy | Kappa |
| --- | --- | --- | --- |
| 3 | 2 | 0.9025617 | 0.8019504 |

### 5.6 Stochastic Gradient Boosting

This model works with a set of decision trees, in which sequentially one tree learns from the preceding one, thus improving the prediction. It uses random sampling of observations without replacement in the training data. This method consists of 4 parameters:

- *shrinkage*: It controls the influence of each model on the final prediction. The values of shrinkage = c(0.001, 0.01, 0.1) were used in training.
- *interaction*.*depth*: It monitors the number of divisions the tree has. The values of interaction.depth = c(1, 2) were selected.
- *n*.*minobsinnode*: It sets a minimum number of observations for the node to be split. In this training a n.minobsinnode = c(2, 5, 15) was used.
- *n*.*trees*: Number of models used in the process. The selected values were n.trees = c(500, 1000, 2000).

Final result taking into account the best Accuracy:

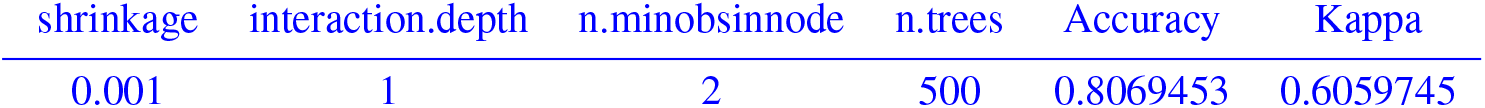

| shrinkage | interaction.depth | n.minobsinnode | n.trees | Accuracy | Kappa |
| --- | --- | --- | --- | --- | --- |
| 0.001 | 1 | 2 | 500 | 0.8069453 | 0.6059745 |

### 5.7 Neural Network

Neural networks are designed with groups of interconnected nodes. Each neural network has 3 types of neurons: input, hidden layer and output. The neurons are interconnected with each other and the strength of this connection is called “weight”. It has the advantage of working well with non-linear relationships between variables and can handle large data diligently. This model has 2 hyperparameters, on the one hand **“size”** which tells us the number of neurons in the hidden layer and **“decay”**, the parameter in charge of regularization during training. In this case we used: size = c(1,3,5) and decay = c(0e+00, 1e-01, 1e-04). Final result and representative graph in Figure 5.

| size | decay | Accuracy | Kappa |
| --- | --- | --- | --- |
| 1 | 0e+00 | 0.8415314 | 0.6814546 |

**Figure 4:**
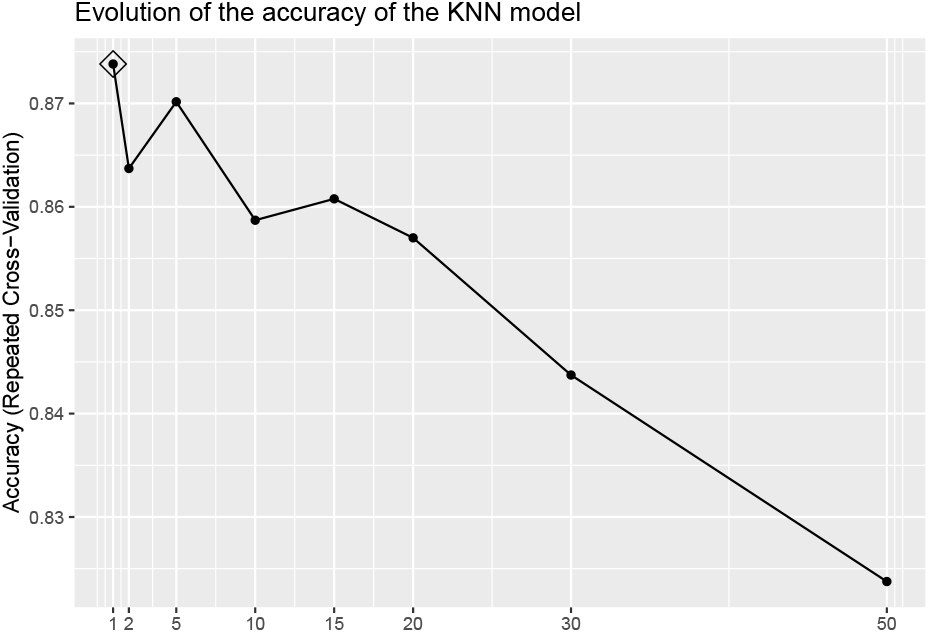
Graphical representation of the training KNN with the values de k used.

**Figure 5:**
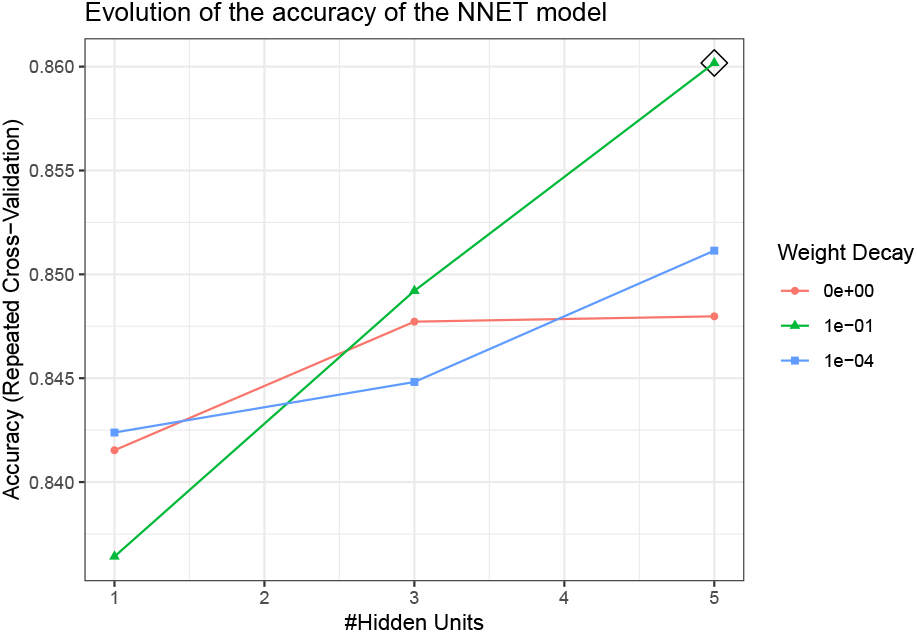
NNET model training.

## 6 Evaluation of the results of all the models

### 6.1 Accuracy and average Kappa of each model

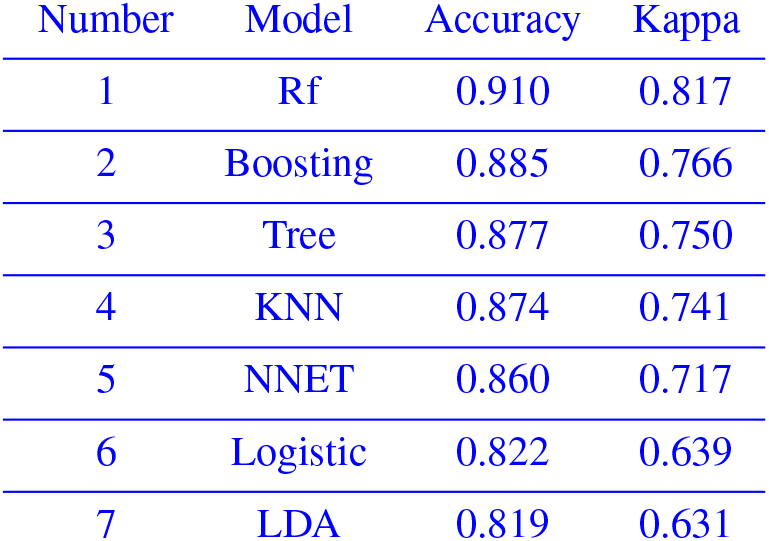

| Number | Model | Accuracy | Kappa |
| --- | --- | --- | --- |
| 1 | Rf | 0.910 | 0.817 |
| 2 | Boosting | 0.885 | 0.766 |
| 3 | Tree | 0.877 | 0.750 |
| 4 | KNN | 0.874 | 0.741 |
| 5 | NNET | 0.860 | 0.717 |
| 6 | Logistic | 0.822 | 0.639 |
| 7 | LDA | 0.819 | 0.631 |

All models exceed the baseline level (0.62) marked on the Figure 6 as a dashed line.

**Figure 6:**
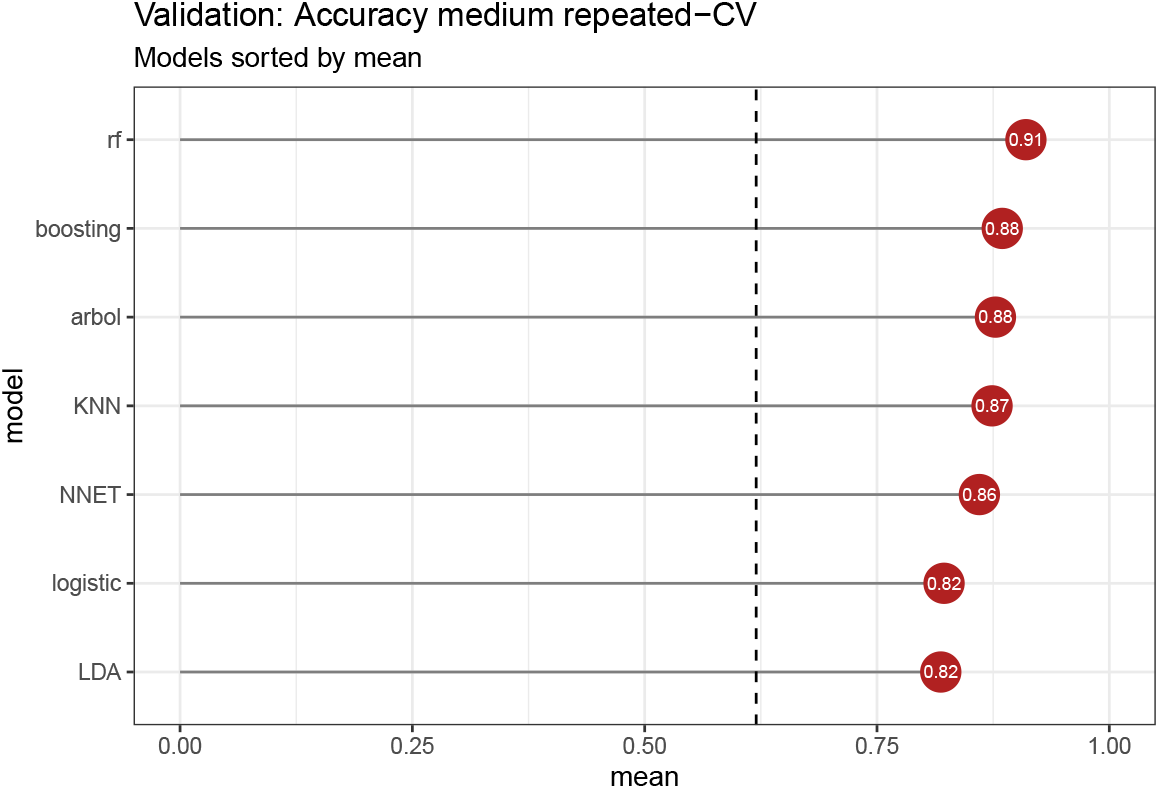
Final results of the training of the different models.

The **Random Forest** model is the one that achieved the best result (Figures 6 and 7). However, we need to compare the variances to determine if the differences are significant (Figure 8).

**Figure 7:**
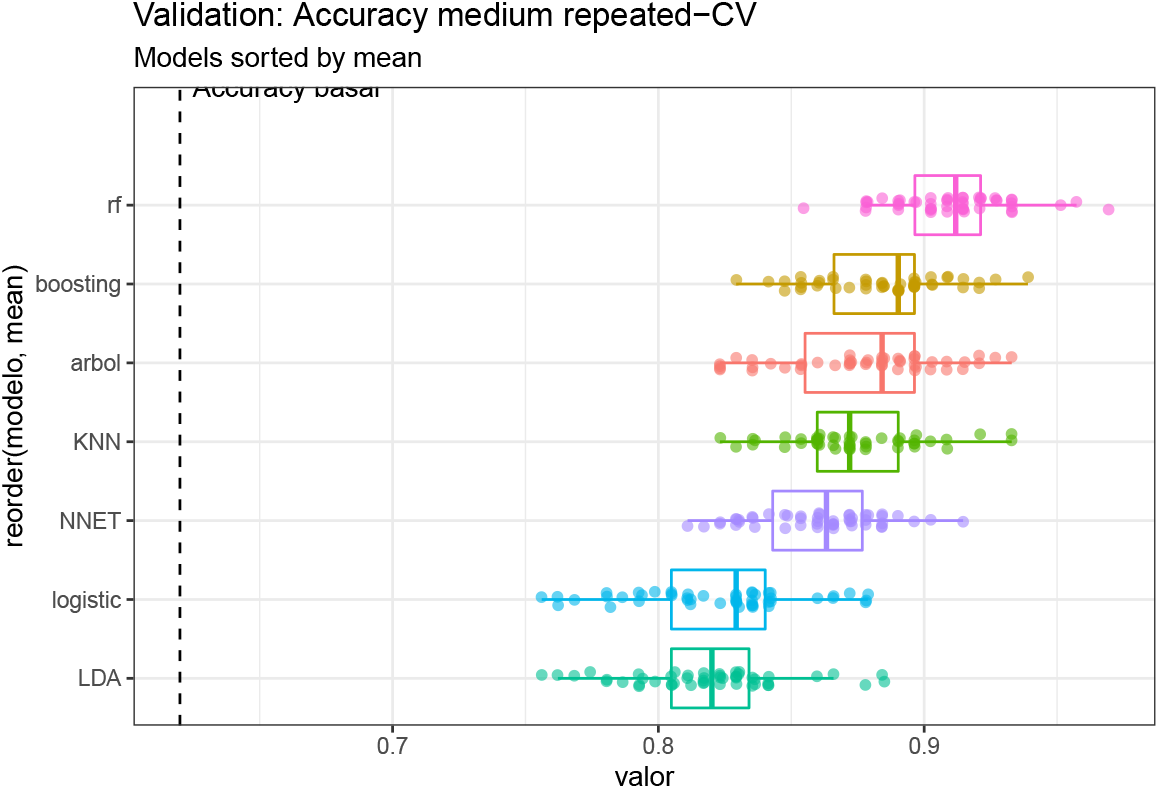
Boxplots of final model results.

**Figure 8:**
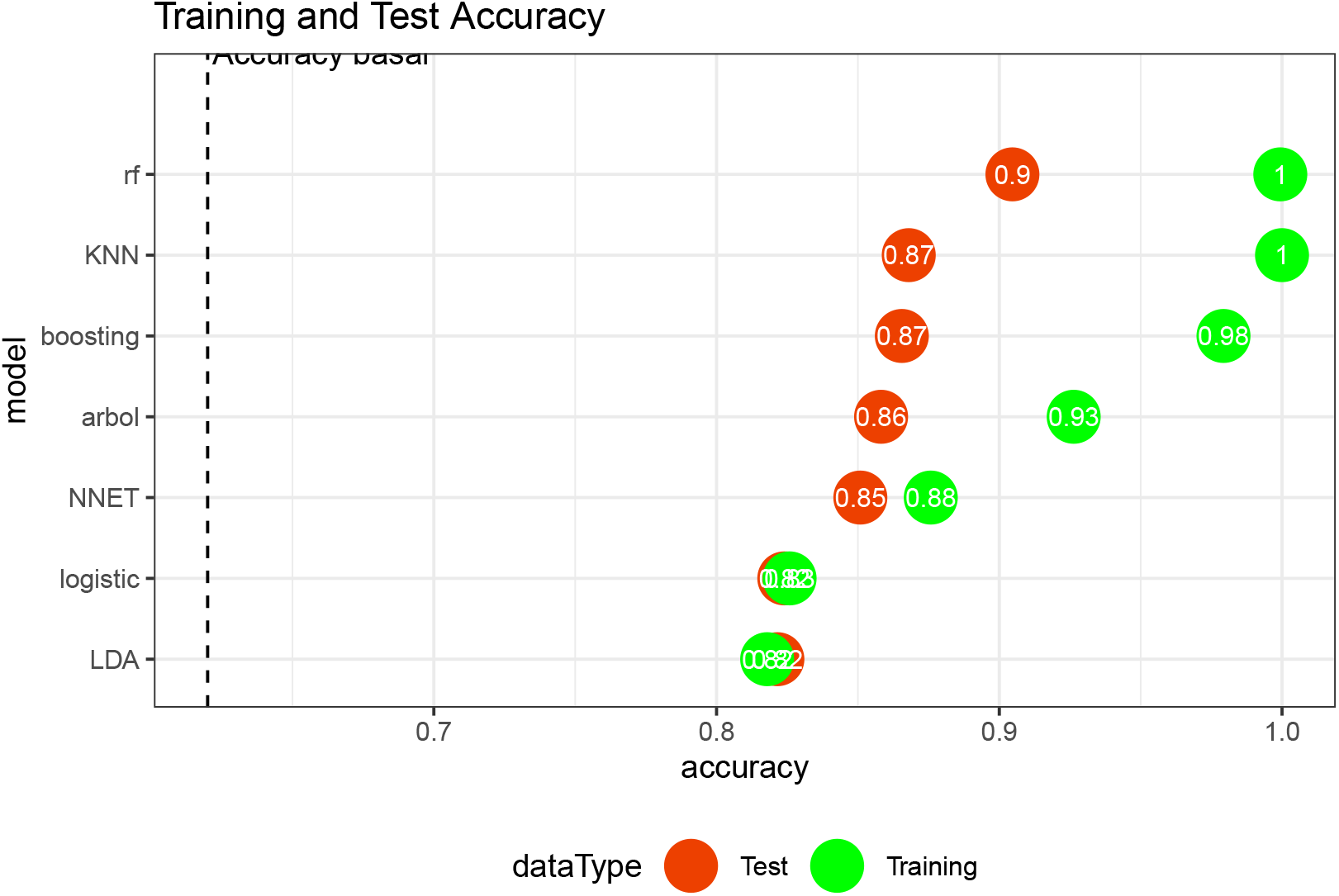
Results of applying the different models to the training and test data.

**Figure 9:**
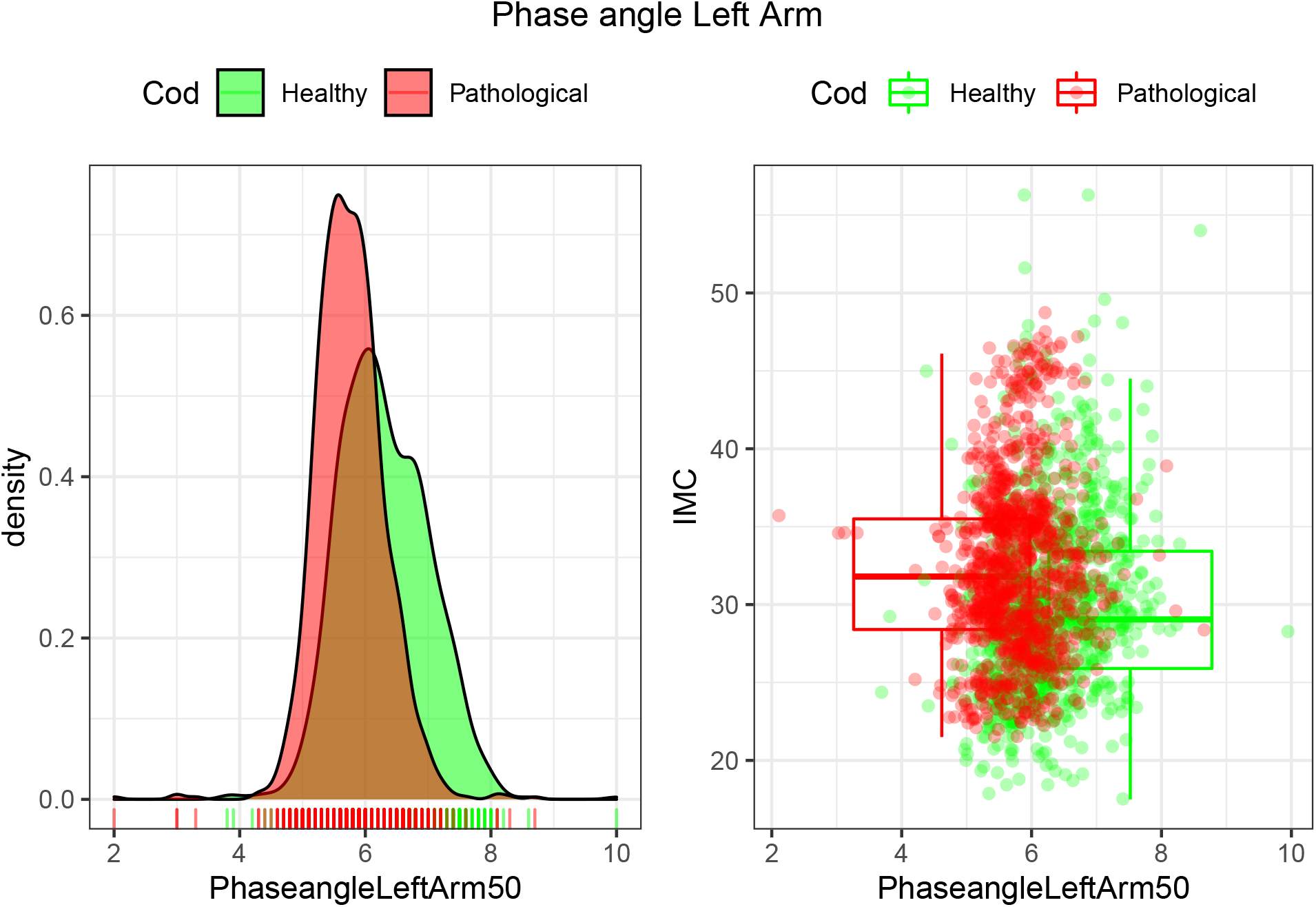
Density and IMC of the two groups based on the phase angle of the left arm with frequencies of 50 kHz.

**Figure 10:**
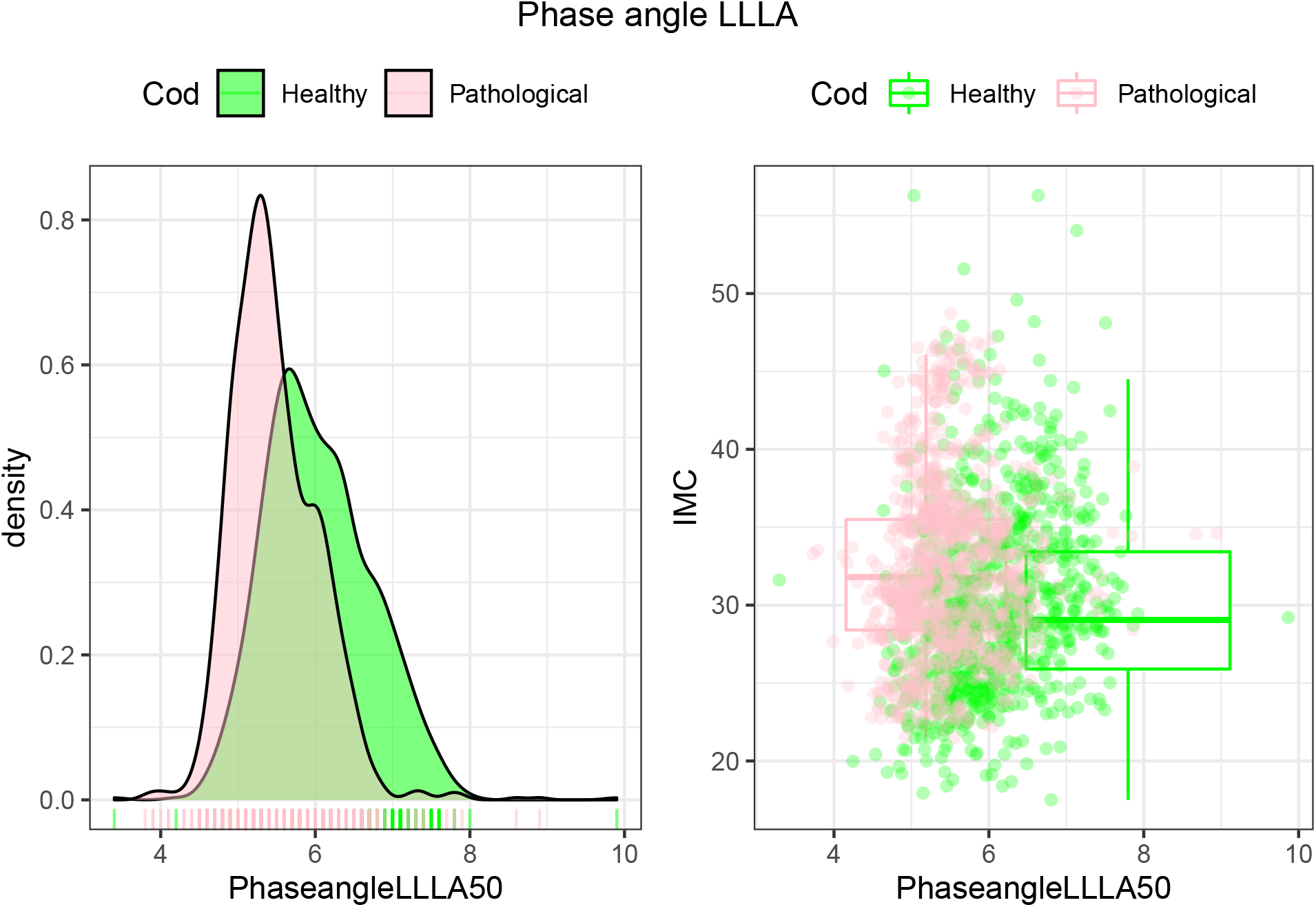
Density and IMC of the two groups based on the phase angle between the left leg and left arm with frequencies of 50 kHz.

**Figure 11:**
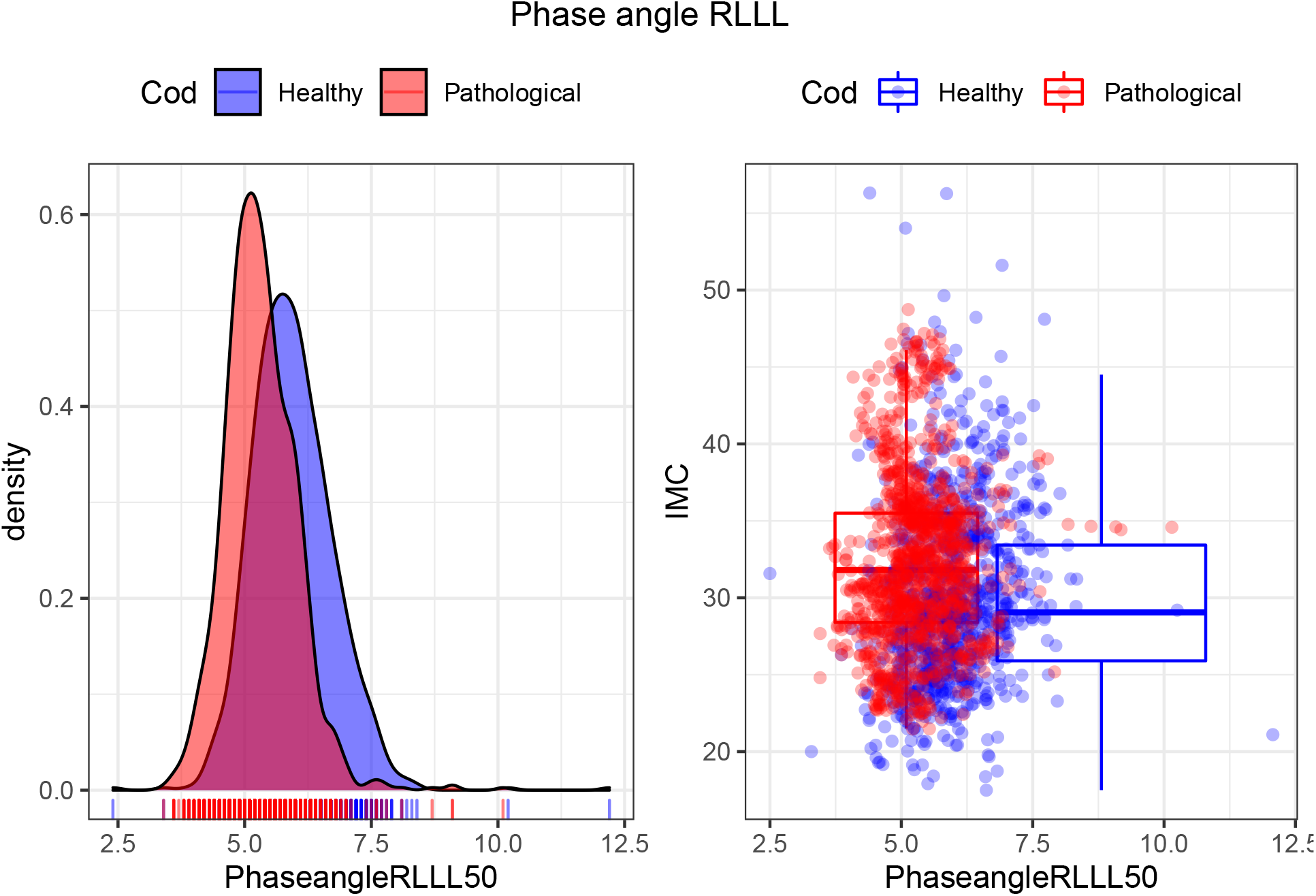
Density and IMC of the two groups based on the phase angle between the right leg and left leg with frequencies of 50 kHz.

**Figure 12:**
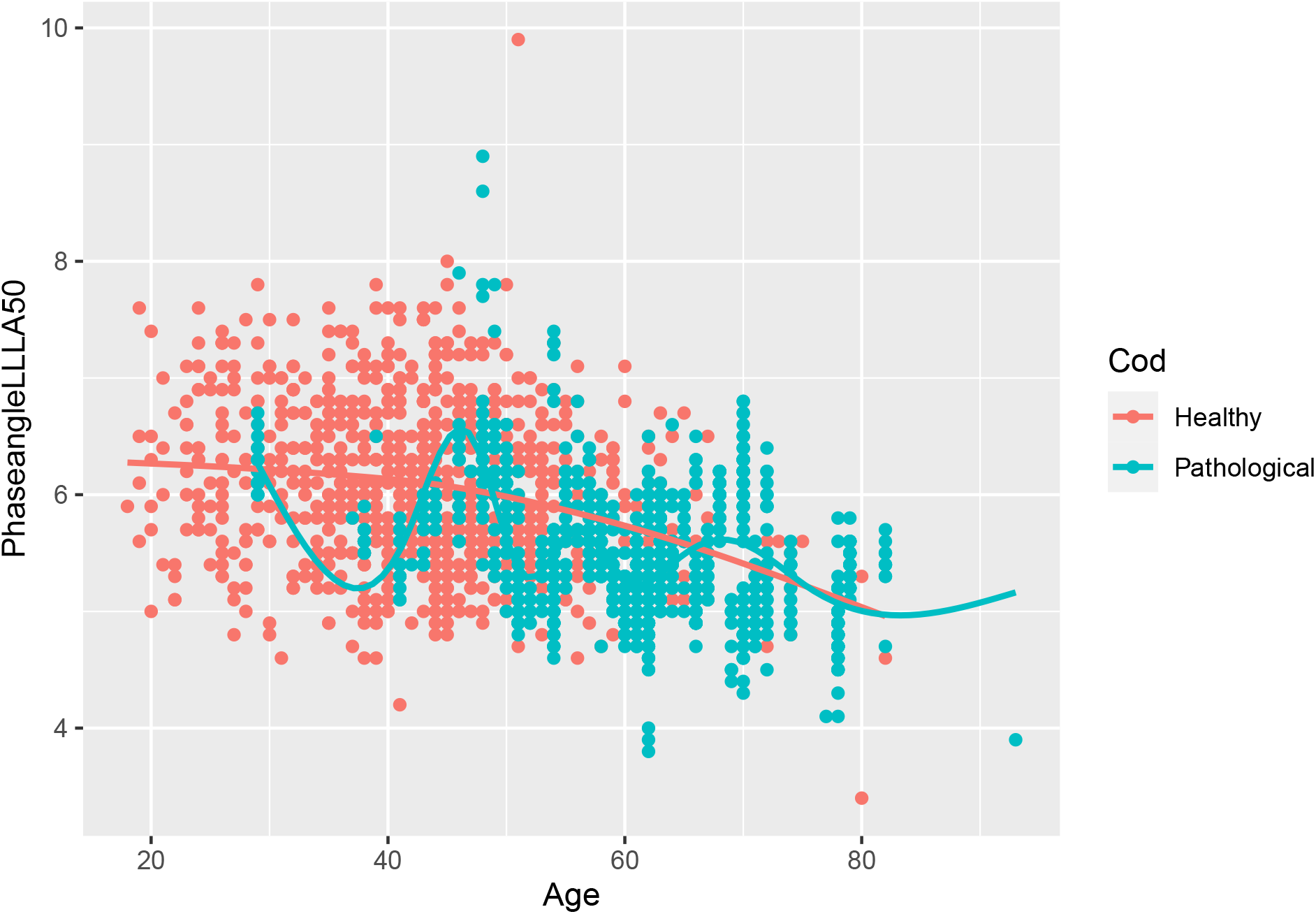
Graphical representation of the phase angle between left leg and left arm according to age in the different groups with frequencies of 50 kHz.

**Figure 13:**
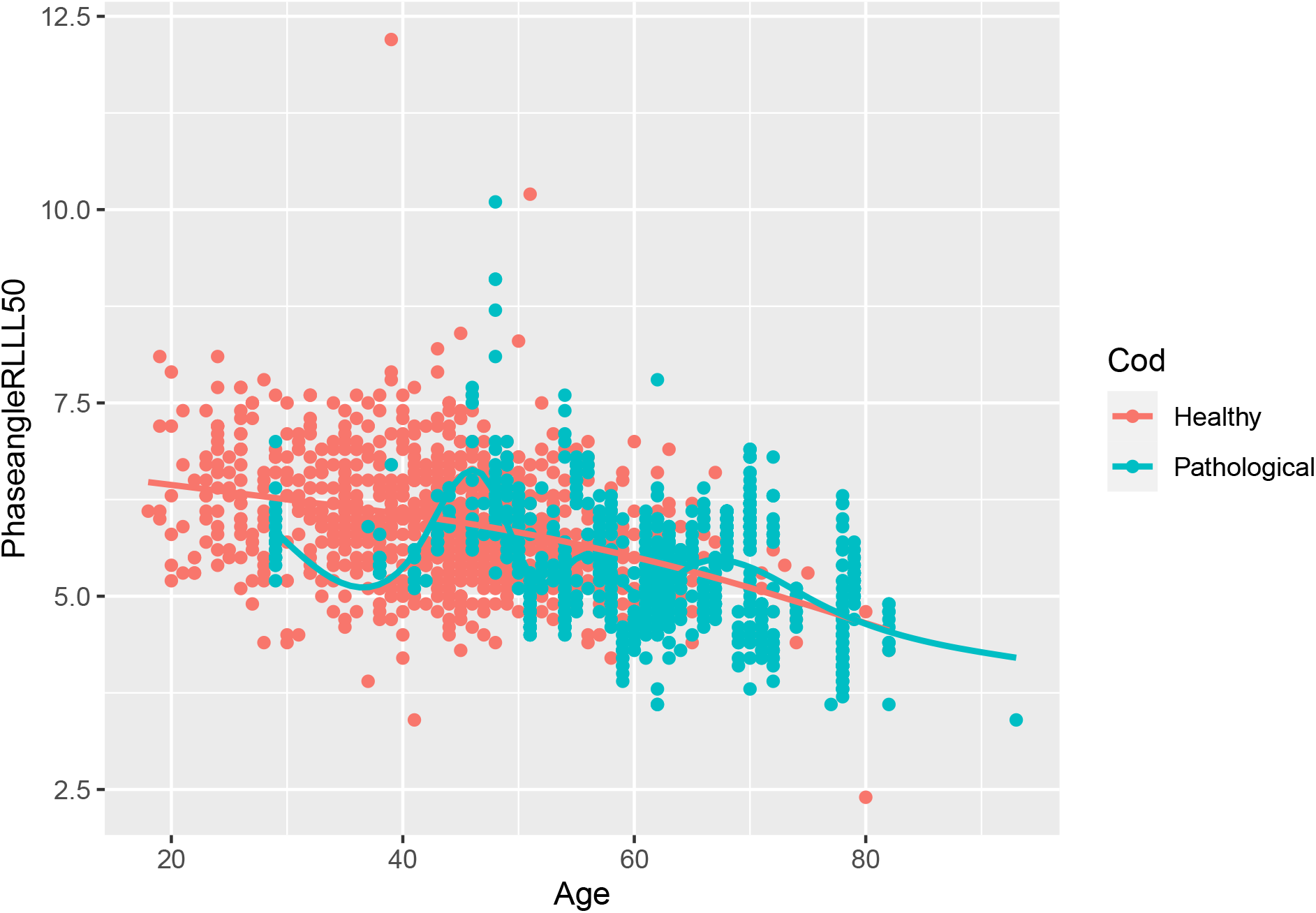
Graphical representation of the phase angle between right leg and left leg according to age in the different groups with frequencies of 50 kHz.

### 6.2 Differences between the training and test set according to the predicted results

When training the data test with the different models, accuracy values higher 82 % were reported:

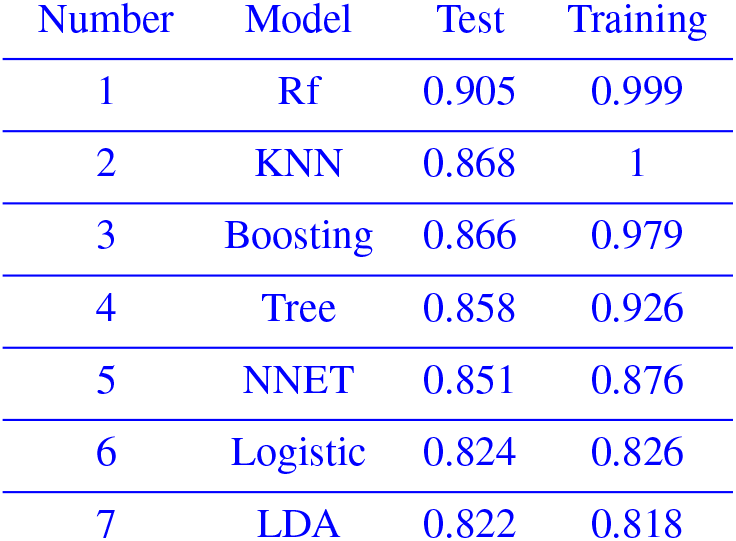

## 7 Discussion

Cancer is not one but a variety of complex diseases whose cellular components are derived from normal cells. Abnormalities of their membranes as well as cell junctions, increased lactate production and the appearance of novel antigens clearly mark the differences between cancer cells and the rest of the cellular pool. These modifications cause changes in the electrical properties of the tissue [68], anticipating even the state of cachexia [63, 62]. This means that alterations in the physiology of the organism’s tissue components are reflected in their electrical properties [67].

The main objective of this study was to use multifrequency bioimpedance analysis with machine learning algorithms in order to predict a high predisposition to neoplastic degeneration based on tissue electrical changes. Machine learning is a discipline in which algorithms and models are applied to a set of data and in this case, the **Random Forest** algorithm reported the best results (according to the accuracy metric) with very good performance. The result achieved in the test was 90 % correct (Figure 8). This model works very well with large databases and is able to handle and catalog numerous predictors [49].

In this study, it is reported that the mathematical algorithms used in machine learning and bioimpedance data are very useful tools to predict an alteration in body homeostasis, a risk situation that may favor the development of neoplastic pathologies or even to predict relapses in patients after neoplastic treatment. There is no doubt that this work has limitations, since a larger sample of people with neoplasias would be needed, as well as a differentiation of them in their different types, stages and treatments received.

## 8 Conclusion

The machine learning-bioimpedance duo is a useful screening tool to detect patients at high risk of neoplastic degeneration, allowing early preventive and therapeutic measures to be used if appropriate.

## Data Availability

All data produced in the present study are available upon reasonable request to the authors.

## Supplementary material

### Variables used to train the neural network

- **Cod**: Classification variable defining the state of health of the individual (pathological or not).
- **Sex**
- **Age**
- **Weight (kg)**
- **ACT**: Total Body Water
- **Fat**: Total fat
- **IMC (BMI)**: Body Mass Index.
- The calculation was based on the Quetelet body mass index formula [19]:

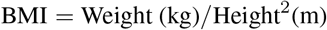

- **Visceral**: Visceral Fat
- **ImpedanceArmLeft**: Impedance Left Arm
- **ImpedanceArmRight**: Impedance Right Arm
- **ImpedanceLegLeft**: Impedance Left Leg
- **ImpedanceLegRight**: Impedance Right Leg
- **PhaseangleLeftLeg50**: Phase angle Left Leg 50 kHz
- **PhaseangleRightArm50**: Phase angle Right Arm 50 kHz
- **PhaseangleLeftArm50**: Phase angle Left Arm 50 kHz
- **PhaseangleLL50**: Phase angle LL 50 kHz
- **PhaseangleRLLL50**: Phase angle RLLL 50 kHz

### Frequency of different types of tumors

As can be seen in Table 1, the malignant tumors with the highest number of cases in this study were breast, thyroid and uterus.

### Training phases of the different models

- Study and preparation of the data.
- Normalization of numerical variables.
- Creation of training and test data.
- Application of machine learning algorithms to the training data.
- Evaluation of the results of the different models.
- Prediction

### Analysis of variance of the models

To determine the most efficient machine learning method, we should not only rely on the metrics, but it is necessary to analyze the variance to establish if there is a clear difference between the different models. In Table 2 we see that the difference in the results of the different models is significant at a significance level of *α*= 0.05.

**Table 2.**

| Model A | Model A | p value |
| --- | --- | --- |
| Boosting | Tree | 1.158947e-01 |
| KNN | Tree | 6.302123e-01 |
| KNN | Boosting | 2.196969e-02 |
| LDA | Tree | 1.998722e-08 |
| LDA | Boosting | 1.998722e-08 |
| LDA | KNN | 4.122123e-08 |
| Logistic | Tree | 1.998722e-08 |
| Logistic | Boosting | 1.998722e-08 |
| Logistic | KNN | 4.890528e-08 |
| Logistic | LDA | 1.158947e-01 |
| NNET | Tree | 6.910109e-04 |
| NNET | Boosting | 1.225374e-06 |
| NNET | KNN | 1.019842e-02 |
| NNET | LDA | 1.998722e-08 |
| NNET | Logistic | 2.451687e-08 |
| Rf | Tree | 8.395390e-08 |
| Rf | Boosting | 8.243194e-08 |
| Rf | KNN | 4.078512e-08 |
| Rf | LDA | 1.581434e-08 |
| Rf | Logistic | 1.581434e-08 |
| Rf | NNET | 1.581434e-08 |

To complete the analysis, we compared only the Random Forest (RF) model with the rest of the models, finding that the difference between them is still significant (Table 3).

**Table 3.**

| Model A | Model B | p value |
| --- | --- | --- |
| Rf | Tree | 8.395390e-08 |
| Rf | Boosting | 8.243194e-08 |
| Rf | KNN | 4.078512e-08 |
| Rf | LDA | 1.581434e-08 |
| Rf | Logistic | 1.581434e-08 |
| Rf | NNET | 1.581434e-08 |

### Graphical representation of the exploratory analysis of the data

